# Preterm birth, stillbirth, and early neonatal mortality during the Danish COVID-19 lockdown

**DOI:** 10.1101/2021.06.09.21258622

**Authors:** Paula L Hedley, Gitte Hedermann, Christian M Hagen, Marie Bækvad-Hansen, Henrik Hjalgrim, Klaus Rostgaard, Anna D Laksafoss, Steen Hoffmann, Jørgen Skov Jensen, Morten Breindahl, Mads Melbye, Anders Hviid, David M Hougaard, Lone Krebs, Ulrik Lausten-Thomsen, Michael Christiansen

## Abstract

**Importance:** Using provisional or opportunistic data, three nationwide studies (The Netherlands, USA, and Denmark) have identified a reduction in preterm or extremely preterm births during periods of COVID-19 restrictions. However, these findings have been challenged as none of the studies accounted for perinatal deaths.

**Objective:** To determine whether the reduction in extremely preterm births, observed in Denmark during the COVID-19 lockdown, could be a result of an increase in number of perinatal deaths; and to assess the impact of extended COVID-19 restrictions on the prevalence of preterm birth and perinatal death.

**Design:** The study is a nationwide Danish register-based prevalence proportion study using detailed data to capture all births (induced abortions were excluded) throughout Denmark. We assessed the prevalence of stillbirth, preterm birth, and early neonatal death.

**Setting:** Population-based study

**Participants:** All singleton pregnancies delivered in Denmark, between February 27, and September 30, 2015-2020. COVID-19 lockdown was studied in 31,164 births and the extended period of COVID-19 restrictions in 214,862 births.

**Exposure:** COVID-19 restrictions broadly (February 27, – September 30, 2020) or COVID-19 lockdown specifically (March 12, – April 14, 2020).

**Main Outcome:** Prevalence of preterm births, stillbirths and early neonatal deaths across the periods under study.

**Results:** The extremely preterm birth rate was reduced (OR 0.27, 95% CI 0.07 to 0.86) during the strict lockdown period, while perinatal mortality was not significantly different. During the extended period of restrictions, the extremely preterm birth rate was marginally reduced, and a significant reduction in the stillbirth rate (OR 0.69, 0.50 to 0.95) was observed. No changes in early neonatal mortality rates were found.

**Conclusion and Relevance:** Stillbirth and extremely preterm birth rates were reduced in Denmark during the period of COVID-19 restrictions and lockdown, respectively, suggesting that aspects of these containment and control measures confer an element of protection. The present observational study does not allow for causal inference; however, the results support design of studies to ascertain whether behavioural or social changes for pregnant women may improve pregnancy outcomes.

**Funding:** None

**Key points:** *Question:* Can changes in stillbirth and early neonatal mortality rates during the COVID-19 lockdown explain the reduction in extremely preterm birth seen in Denmark?

*Findings:* In this nationwide register-based study that included data pertaining to 245,999 pregnancies, the statistically significant reduction in extremely preterm birth during the COVID-19 lockdown was confirmed. The stillbirth and early neonatal mortality rates were unchanged during lockdown, while the stillbirth rate was reduced over the extended period of COVID-19 restrictions.

*Meaning:* The reduction in extremely preterm births seen during the COVID-19 lockdown in Denmark is not a result of increased stillbirth or early neonatal death.

## Introduction

COVID-19 containment and control policies were implemented worldwide in response to the SARS-Cov-2 pandemic. The preventative and protective measures placed on communities in order to reduce viral transmission created an opportunistic experiment^1^ which may add to our future understanding of what causes preterm birth (PTB).^2^ A Danish nationwide study first described a dramatic reduction in extremely PTBs (xPTB) during the strict lockdown period.^3^ Following this finding, similar reductions in PTBs were reported from the Netherlands,^1^ Japan,^4^ Italy,^5^ Tennessee,^6^ New York,^7^ Israel,^8^ and the USA^9^ and a report from Ireland described a reduced proportion of very low birth weight babies.^10^ However, smaller studies from California,^11^ Philadelphia,^12^ Israel,^13^ Spain,^14^ and London,^15^ as well as a recent nationwide Swedish study^16^ could not confirm these findings.

A recent meta-analysis indicated that during the COVID-19 pandemic high-income countries generally saw a reduction in PTBs and stillbirths, while low-to -middle-income countries (LMIC) saw increases.^17^ Notably there are very few reports from LMIC. The conflicting findings can in part be explained by health system inefficiencies and/or an inability to deal adequately with the pandemic.^17^ In Nepal, for instance, the number of women giving birth in institutions dropped precipitously.^18^ These regional differences are further supported by a prepublication reporting data from 17 countries,^19^ where the xPTB rate was reduced between 11 % and 22 % in the European region, and increased by 48 % in China and India. Furthermore, some studies have reported elevated stillbirth rates^5, 13, 15^ and a study from Nepal reported an increase in both PTBs and stillbirths during their lockdown periods.^18^

Stillbirths and early neonatal mortality rates were not assessed in the three nationwide studies from The Netherlands, USA, and Denmark^1, 3, 9^. Since PTB rates could be reduced in response to increasing perinatal mortality rates^20^ there is a need to assess live birth rates alongside perinatal mortality rates (stillbirth and early neonatal death) during the different periods studied. Accordingly, this study aimed to use data pertaining to all Danish pregnancies and births, captured in the extensive nationwide electronic registers to study the prevalence of PTB, stillbirth, and early neonatal mortality from singleton pregnancies during the first strict lockdown period in Denmark and during the continuous period of COVID-19 restrictions. Furthermore, we describe the stringency of Danish COVID-19 policies and the timeline of their implementation in order to contextualise our findings.

## Methods

### Study design and participants

The study is a retrospective register-based, nationwide study comparing the prevalence’s of PTB and perinatal death (stillbirth, and early neonatal death defined as death within the first seven days of life) in the Danish COVID-19 lockdown period with the same calendar periods in the preceding five years. All singleton births, born with a gestational age ≥ 22+0 weeks, registered in these periods, were included and the outcome, i.e., stillbirth, live birth at specific gestational ages, as well as death within the early neonatal period (first seven days of life) – were registered. Induced abortions were excluded.

### Data sources

Participants were identified using the extensive population-based registers available in Denmark,^21^ specifically The Danish Civil Registration System,^22^ the Medical Birth Registry,^23^ and The Danish National Patient Registry (LPR)^24^ (LPR2 and, from February 2019, LPR3). Country specific information about stringency and temporality of COVID-19 lockdown measures was obtained from the Oxford COVID-19 Government Response Tracker^25^. Mobility information from cell phone registrations were obtained from Google COVID-19 Community Mobility Reports.^26^

### Study period and outcomes

Births taking place between March 12, and April 14, 2020 constituted the group exposed to strict lockdown measures, and births between February 27, and September 30, 2020 constituted the broader group exposed to COVID-19 restrictions. The unexposed, control group, was the aggregated deliveries in the calendar periods February 27, – September 30, and March 12, – April 14, 2015-2019, respectively.

### Statistical analysis

Prevalences were compared using Fisher’s exact test and proportionality test. Data analysis was performed using R version 4.0.3.

### Ethics approval

The study was conducted according to Danish legislation and guidelines for register research and was approved by the Data Protection Agency officer at Statens Serum Institut (No: 20/04753)

## Results

### The Danish lockdown

A strict lockdown was in effect from March 12 to April 14, 2020, followed by varying COVID-19 restrictions from April 15 to the end of this study, September 30, 2020. The chronological development of the official Danish response to the COVID-19 pandemic is summarised in Figure 1, and the instituted measures, as well as behavioural effects, are described in detail in the eAppendix and eFigures 1A and 1B in the Supplement.

**Figure 1.**
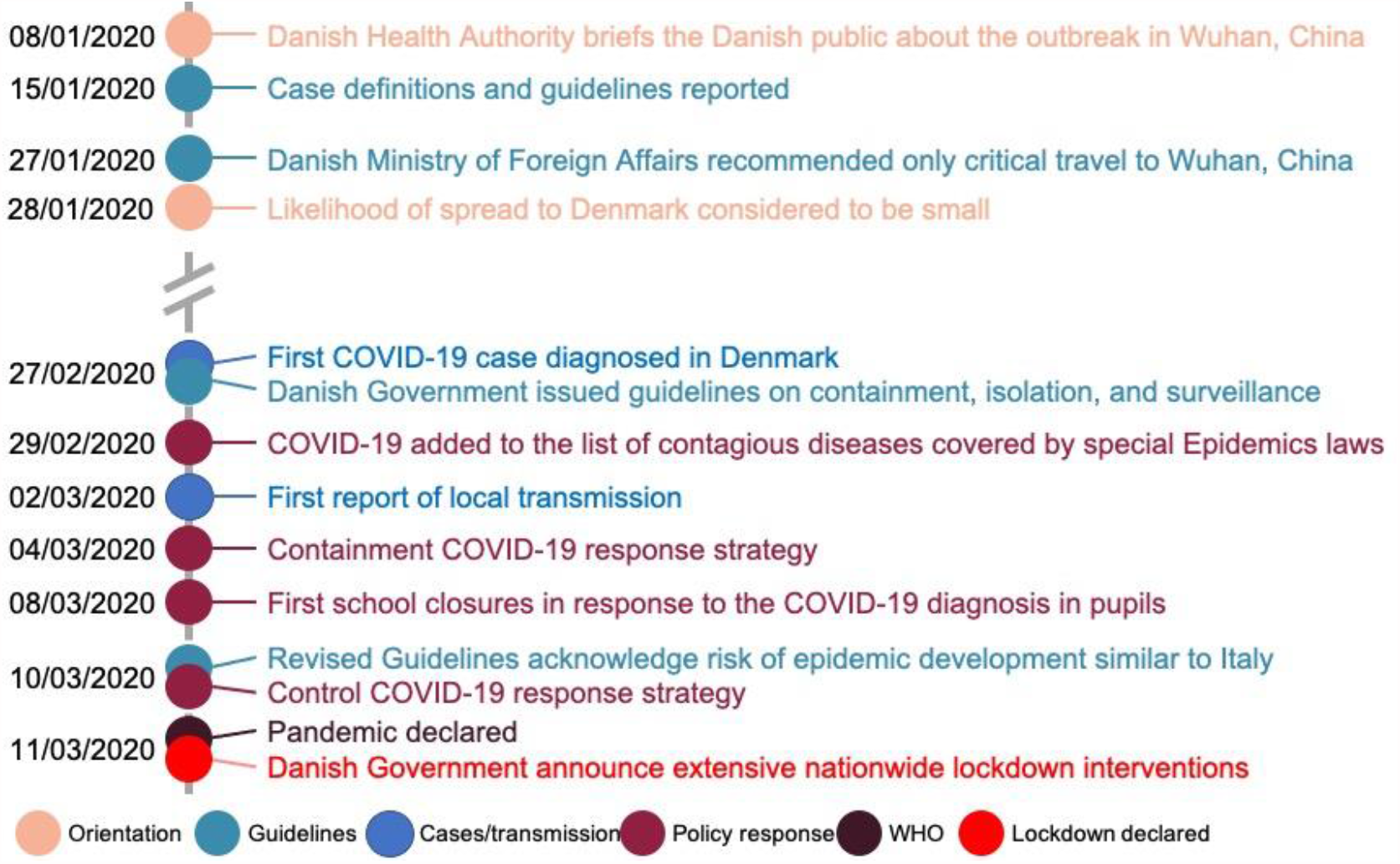
The timeline of COVID-19 events in Denmark leading up to the lockdown. Different categories of events (orienting the public = Orientation, published guidelines = Guidelines, local cases and local transmission = Cases/transmission, policy response = Policy response, the WHO declaration of a pandemic = WHO, the declaration of the lockdown = Lockdown declaration) are indicated by different colours. Data summarised from.^31, 54, 55^

### Birth and Mortality Rates During The Strict lockdown

A total of 31,164 pregnancies with gestational age ≥ 22+0 weeks were registered in Denmark during the period March 12, – April 14, in 2015 – 2020. No differences in birth rates, or perinatal mortality rates were found between the year of 2020 and preceding years 2015 – 2019 (Table 1). However, the xPTB rate was significantly reduced (OR 0.27, 95% CI 0.07 – 0.86) while the other PTB groups were not changed (Table 2). There were no differences in gestational age dependent stillbirth rates, perinatal mortality and neonatal mortality rates (Table 2).

**Table 1.** Number of births, stillbirths, and mortality rates in the lockdown (March 12 – April 14) and extended period of COVID-19 restrictions (February 27 – September 30).

|  | 2020 | 2015 - 2019 | 2020 vs 2015-2019 |
| --- | --- | --- | --- |
| **Strict lockdown (March 12 – April 14)** |  |  |  |
| Births | N | Mean (SD) |  |
| Total births | 5,013 | 5,230.2 (207.4) |  |
| Total live births | 4,999 | 5,215.0 (205.4) |  |
| % male | 51.8 | 51.3 (0.62) |  |
| Mortality and mortality rates | N (%) | Mean (SD) | OR (95% CI) |
| Perinatal mortality | 19 (0.38) | 23.2 (3.4) | 0.86 (0.44, 1.66) |
| Stillbirths | 14 (0.28) | 15.4 (6.1) | 0.97 (0.43, 2.17) |
| Early Neonatal mortality ($\leq 7$ days) | 5 (0.10) | 7.8 (2.9) | 0.65 (0.17, 2.26) |
| Very Early mortality (< 24 hrs) | 3 (0.10) | 5.6 (1.5) | 0.52 (0.08, 2.44) |
| **Period of COVID-19 restrictions (February 27 – September 30)** |  |  |  |
| Births | N | Mean (SD) |  |
| Total births | 35,394 | 35,893.6 (1,053.9) |  |
| Total live births | 35,326 | 35,793.4 (1,048.3) |  |
| % male | 51.4 | 51.5 (0.21) |  |
| Mortality and mortality rates | N (%) | Mean (SD) | OR (95% CI) |
| Perinatal mortality | 111 (0.31) | 148.2 (25.8) | 0.76 (0.29, 0.98) |
| Stillbirths | 68 (0.19) | 100.2 (14.9) | 0.69 (0.50, 0.95) |
| Early Neonatal mortality ($\leq 7$ days) | 43 (0.12) | 48.0 (11.8) | 0.91 (0.59, 1.40) |
| Very Early mortality (< 24 hrs) | 22 (0.06) | 28.0 (8.7) | 0.80 (0.43, 1.44) |

**Table 2.** Live births and stillbirths as a function of gestational age at birth in the lockdown (March 12 – April 14) and extended period of COVID-19 restrictions (February 27 – September 30).

|  | GA | 2020 | 2015-2019 | 2020 vs 2015 - 2019 |
| --- | --- | --- | --- | --- |
| **Lockdown (March 12 – April 14)** |  |  |  |  |
|  | Weeks +days | N (%) | Mean (SD) | OR (95% CI) |
| Live births |  |  |  |  |
| Extremely preterm | ≤27+6 | 4 (0.08) | 15.0 (3.2) | 0.27 (0.07, 0.86) |
| Very preterm | 28+0-31+6 | 18 (0.36) | 23.4 (1.6) | 0.80 (0.41, 1.56) |
| Moderate preterm | 32+0-36+6 | 202 (4.03) | 217.4 (7.3) | 0.96 (0.78, 1.17) |
| Term | 37+0-41+6 | 4,731 (94.37) | 4,835.0 (206.1) | 1.13 (0.95, 1.34) |
| Late term | ≥42+0 | 33 (0.66) | 40.6 (3.8) | 0.83 (0.51, 1.34) |
| Missing GA |  | 11 (0.22) | 84.0 (39.5) |  |
| Stillbirths |  |  |  |  |
| Extremely preterm | ≤27+6 | 4 (0.08) | 6.2 (3.0) | 0.55 (0.08, 3.28) |
| Very preterm | 28+0-31+6 | 4 (0.08) | 2.6 (1.10) | 1.57 (0.21, 13.43) |
| Moderate preterm | 32+0-36+6 | 2 (0.04) | 2.4 (1.1) | 1.00 (0.06, 15.94) |
| Term | 37+0-41+6 | 4 (0.08) | 3.2 (1.5) | 1.45 (0.19, 12.44) |
| Late term | ≥42+0 | 0 (0.00) | 0.0 (0.0) | 0.00 (0.00, inf) |
| Missing GA |  | 0 (0.00) | 0.0 (0.0) |  |
| **Period of COVID-19 restrictions (February 27 – September 30)** |  |  |  |  |
|  | Weeks +days | N (%) | Mean (SD) | OR (95% CI) |
| Live births |  |  |  |  |
| Extremely preterm | ≤27+6 | 58 (0.16) | 74.6 (4.9) | 0.79 (0.55, 1.12) |
| Very preterm | 28+0-31+6 | 150 (0.43) | 165.8 (6.4) | 0.92 (0.73, 1.15) |
| Moderate preterm | 32+0-36+6 | 1,410 (3.98) | 1,468.6 (45.8) | 0.98 (0.90, 1.05) |
| Term | 37+0-41+6 | 33,338 (94.19) | 33,242 (903.4) | 1.38 (1.29, 1.46) |
| Late term | ≥42+0 | 241 (0.68) | 289.4 (23.4) | 0.85 (0.71, 1.01) |
| Missing GA |  | 129 (0.36) | 550.8 (249.0) |  |
| Stillbirths |  |  |  |  |
| Extremely preterm | ≤27+6 | 22 (0.06) | 33.4 (6.4) | 0.97 (0.47, 1.97) |
| Very preterm | 28+0-31+6 | 9 (0.03) | 11.6 (3.6) | 1.13 (0.39, 3.13) |
| Moderate preterm | 32+0-36+6 | 16 (0.05) | 20.0 (6.3) | 1.23 (0.54, 2.76) |
| Term | 37+0-41+6 | 21 (0.06) | 32.4 (6.1) | 0.95 (0.46, 1.94) |
| Late term | ≥42+0 | 0 (0.00) | 0.2 (na) | 0.00 (0.00, inf) |
| Missing GA |  | 0 (0.00) | 2.6 (1.7) |  |

### Birth and Mortality Rates During The Extended period of COVID-19 restrictions

A total of 214,862 births with gestational age ≥ 22+0 weeks were registered in Denmark during the period February 27 – September 30, 2015-2020. The number of live births in 2020 was similar to the mean number of live birth per years during 2015 – 2019 (Figure 2A), but the number of stillbirths in 2020 was reduced in all months, except April (Figure 2B). The number of extremely preterm live births was reduced, albeit insignificantly, in all months, except May (OR 2.72, 1.40 – 5.09) and June (OR 2.33, 1.05 – 4.83) where they were significantly increased (Figure 2C). The number of extremely preterm stillbirths was uniformly reduced in all months (Figure 2D). The live birth rate, for all PTB groups, was not different in 2020 (Table 2). Furthermore, the proportion of term pregnancies was increased (OR 1.38, 1.29 – 1.46), whereas the proportion of late term pregnancies was reduced (OR 0.85, 0.71 – 1.01) (Table 2). Importantly, the total stillbirth rate was reduced (OR 0.69, 0.50 – 0.95), resulting in a corresponding reduction in perinatal mortality (OR 0.76, 0.20 – 0.98) (Table 1). The very early and early neonatal mortality rates were not different between 2020 and 2015 – 2019 (Table 1).

**Figure 2.**
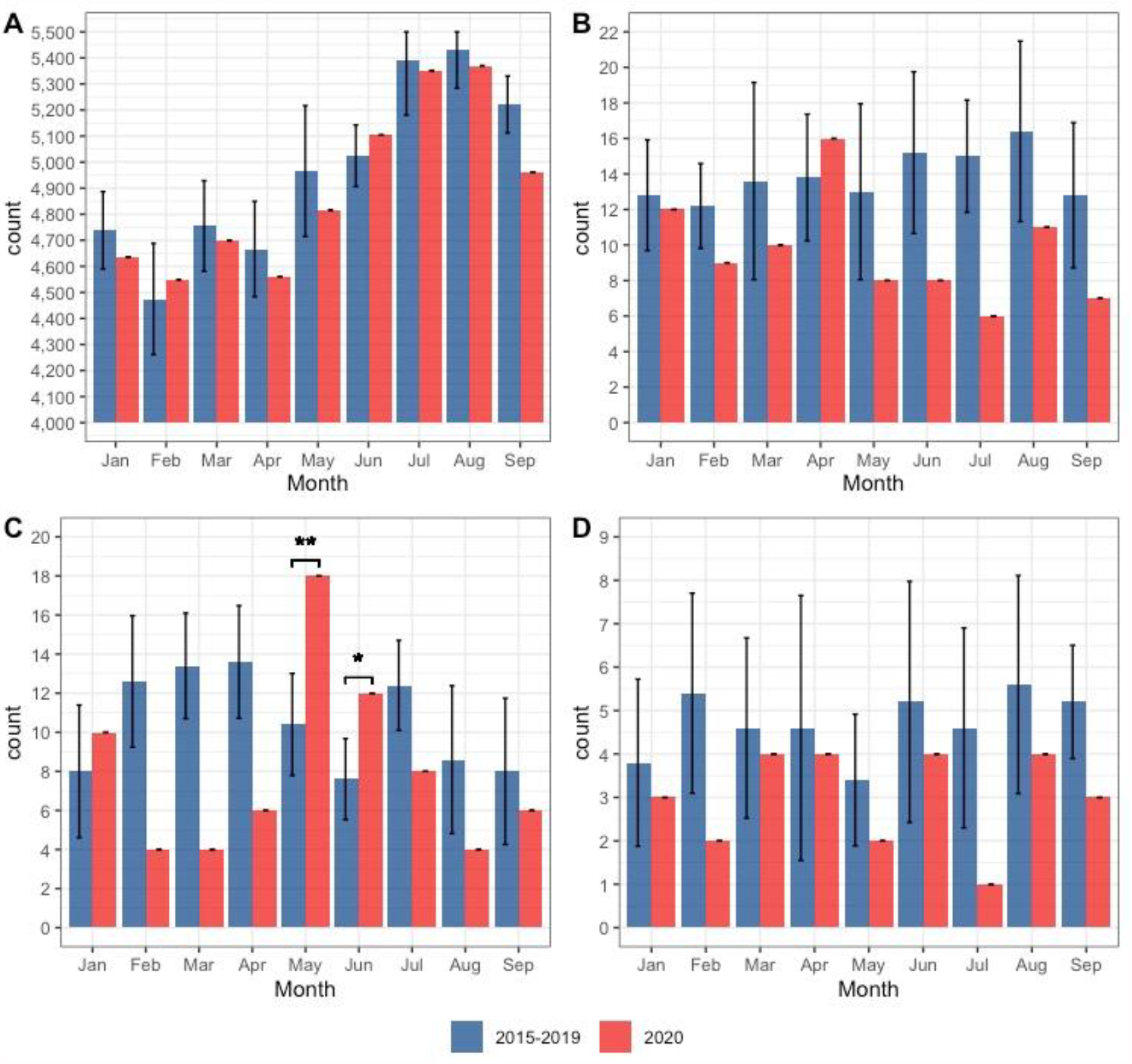
Mean number of live births and stillbirths per month, from January to September. 2015-2019 (blue; error bars represent 1SD) and 2020 (red). **A:** live births (all gestational ages), **B:** stillbirths (all gestational ages, **C:** livebirths (extremely preterm, gestational age 22-28 weeks), and **D:** stillbirths (gestational age 22-28 weeks). Statistically significant differences are indicated by asterisks. * represents p ≤0.05, and ** represents p <0.01.

### Combined Extremely Preterm Live and Still birth Rates

The monthly combined stillbirth and live birth rates for infants delivered with gestational age between 22+0 and 28+0 weeks (extremely preterm), are illustrated in Figure 3. Despite a non-significant increase in May and June, the combined xPTB rate (live and still) during the COVID-19 restriction period was significantly reduced (2.8 per 1000 births compared to 3.8 per 1000 births in 2015-2019) (OR 0.73, 0.59 – 0.96). The increase in May and June is underpinned by an increase in xPTBs (live births) (OR 2.72, 1.40 – 5.09, and OR 2.33, 1.05 – 4.83), respectively) (Figure 2D). May and June are months where restrictions were eased (eFigure 1A) resulting in a marked increase in workplace attendance, public transport use, and visits to retail spaces alongside a reduction in time spent in places of residence (eFigures 1C & 2).

**Figure 3.**
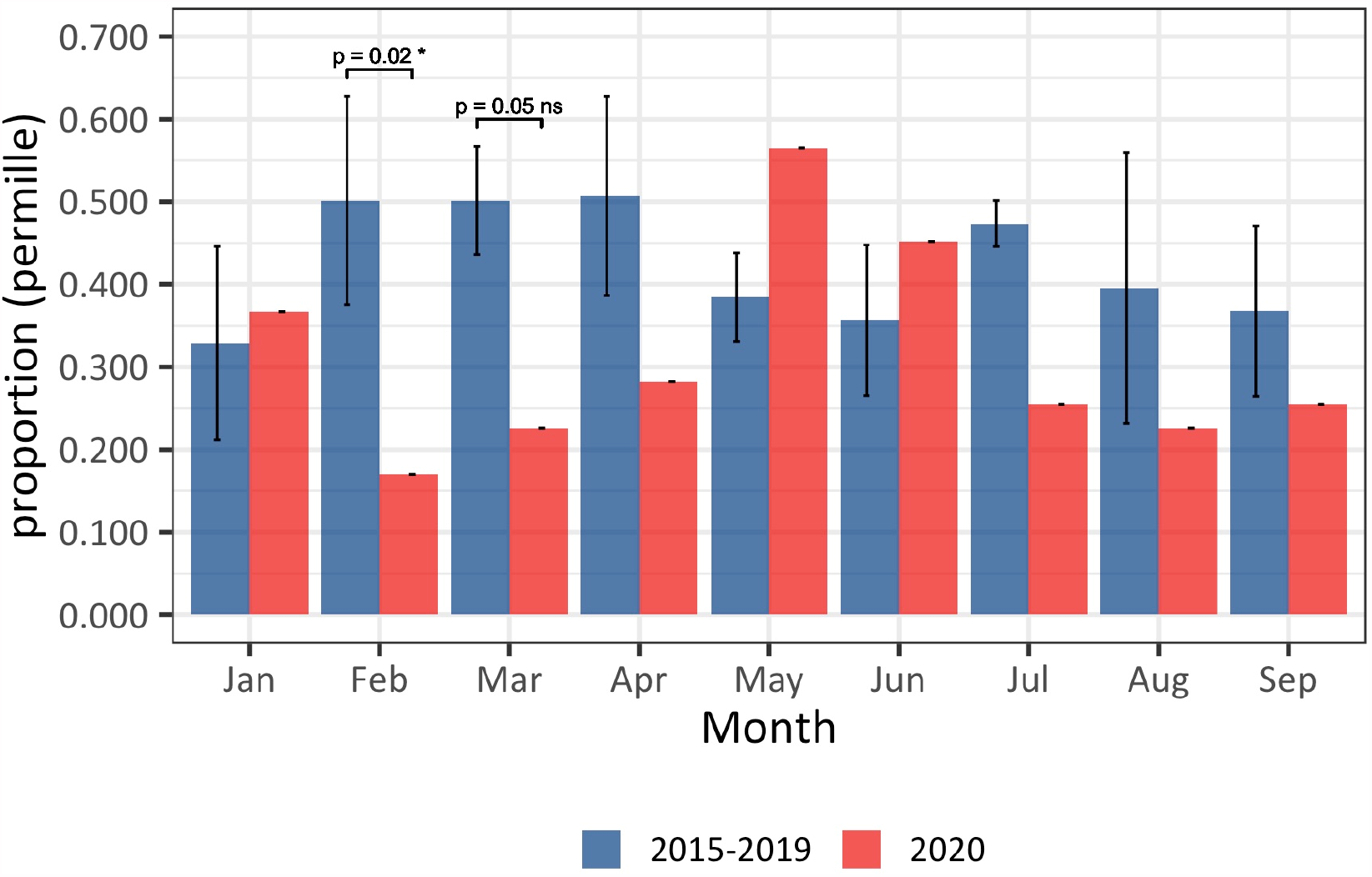
The effect of COVID-19 restrictions on combined extremely preterm stillbirth and live birth rates. Combined rates for 2020 (red columns) are compared to the aggregate rates from the same months in 2015 – 2019 (blue columns; error bars represent 1SD). The sum of the rates is significantly decreased in February and March. Furthermore, the combined rate is decreased in all months except May and June 2020.

## Discussion

Prematurity is a complex and challenging pathophysiological condition^27^ which is the leading cause of death in children under five years old.^28, 29^ The complex and multifaceted aetiology of PTB is linked to a wide range of psychosocial, medical, and environmental factors, but remains only partly understood.^30^ Emerging data suggest that the COVID-19 pandemic and subsequent mitigation measures have provided a natural experiment, through which we can compare PTB and perinatal mortality rates against the human psychosocial and environmental characteristics of pre-COVID-19 and COVID-19 restriction periods.

Using population-based Danish register data, we have documented that the strict lockdown period is associated with a ∼70 % reduction in singleton xPTBs, while no difference was noted for the other PTB groups (Table 2). This result is compatible with the previously reported ∼90 % reduction in xPTBs assessed from the Danish Neonatal Screening Biobank (DNSB),^3^ taking into account that three out of four extremely preterm infants died during the first day of life (very early neonatal death) – precluding their appearance in the DNSB (Table 1). The proportion of very PTBs was slightly, ∼20 %, reduced, but there is no evidence for a protective effect of the lockdown in this group. It is important to note that the reduction in xPTBs during the lockdown period does not appear to be mirrored by an increase in stillbirths (Table 1 & 2).

A reduction in PTB or xPTBs has been observed in three nationwide studies from high income countries (The Netherlands, USA, and Denmark^1, 3, 9^). The stringency of lockdown in all three countries was greater than 70 %, (eFigures 1, 3, and 5 in the Supplement), with clear effects on location behaviour, e.g., workplace attendance reduced around 50 %, (eFigures 1, -6 in the Supplement). A nationwide study conducted in Sweden found no effect on the rate of PTB,^16^ however, the Swedish COVID-19 response was characterized by comparatively lenient lockdown measures, with a stringency of approximately 65 % (eFigure 7 in the Supplement). Mobility data from Sweden reflects these measures with an average reduction of workplace attendance of approximately 30 % (eFigures 7 & 8 in the Supplement). These data suggest that the reduction in xPTBs correlates with the nature and extent of the lockdown, and, in consequence, the behavioural and psychosocial changes associated with an effective lockdown.

The data presented in the current study, covers a broader period of the Danish COVID-19 restrictions, and demonstrates the temporal variation in xPTB rates after the easing of the lockdown (Apr 15 – Sep 30) as illustrated by a marked increase in xPTB rates in May and June (Figure 2C) followed by reduced levels in July, August and September. The net result is that for the whole period with COVID-19 restrictions, the odds ratio of xPTB was reduced, albeit not significantly (Table 2). The months May and June coincide with a reopening of society and relative return to normal activities, (eFigures 1C & 2). For example, the lockdown is characterized by a stable ∼50 % reduction in workplace attendance, during May and June workplace attendance climbs steadily to ∼20 % reduction where, with the exception of vacation period in July, it remains relatively stable. Thus, the periods characterized by high stringency and considerable mobility reduction (lockdown) correlates with a significant reduction in xPTB, whereas the lockdown easing phase (May and June) coincides with a significant increase in xPTB rate.

The stillbirth rate for the whole COVID-19 restriction period (February 27 – September 30) was reduced by ∼30 % (Table 1). The monthly number of total stillbirths (Figure 2B) and stillbirths among extremely preterm pregnancies (Figure 2D) does indicate a uniform reduction in the number of stillbirths in each month during the COVID-19 restriction period. Consequently, the stillbirth rate does not account for the reduction in xPTBs during the Danish lockdown and is itself reduced as a possibly unintended beneficial consequence of the COVID-19 restrictions generally.

The reduction in combined extremely preterm stillbirth and live birth rates is statistically significant in February and March, while the increase in May and June is not (Figure 3). This seems to reflect that the improvement in perinatal health started prior to the lockdown, during which period the Danish government communicated frequently with the public regarding the risk of COVID-19 in Denmark and societal preparedness (Figure 1).^31^

Perinatal mortality was moderately reduced during the lockdown period, and was, as a result of a considerably reduced stillbirth rate, significantly reduced over the extended COVID-19 restriction period (Table 1). Early, and very early, neonatal mortality was not changed during these periods (Table 1). Consequently, despite the strain COVID-19 is expected to place on health care resources,^32^ the observed reduction of xPTB is unlikely to be driven by a change in obstetric policies, formal or otherwise.

One may speculate on the causes of the reduction in xPTBs and stillbirths – one suggestion is a reduction in exposure to harmful pathogens^33^ or changes in genital tract microbiota due to hygiene precautions; massive reductions in the transmission of meningitis^34^ and pertussis,^35^ and an absent influenza season 2020-2021^36^ are consistent with this argument. Furthermore, the number of laboratory confirmed chlamydia cases dropped ∼25 % during the lockdown compared to the 2015-2019 average, followed by an increase in June and July 2020, and a return to the 2015-2019 level in August.^37^ However, the number of people tested for chlamydia during this time is unknown. As a surrogate marker of ascending urogenital infections and microbiota changes, this moderate reduction during the lockdown suggests that a reduction in ascending infections is unlikely to be responsible for the reduction of xPTBs. Exposure to environmental pollutants, particularly air pollution, has been associated with PTB.^38^ However, the Danish lockdown was not associated with major changes in the level of air pollution.^39^

Reduced anxiety and stress may also play a role by influencing the expression of the chaperone protein FKBP51; which increases in response to stress and alters the oestrogen/progesterone ratio thus overcoming the progesterone effect which inhibits parturition.^40, 41^ Two, albeit nearly 30 year old, Danish studies showed an association between anxiety in gestational week 30, but not in week 16, and PTB,^42^ and an association between very early PTB (< 34 weeks) and severe stressful events in mid-pregnancy,^43^ respectively. Furthermore, leisure-time activity during pregnancy has been associated with reduced risk of PTB,^44^ while hard work and hard physical activity during pregnancy could be increase the risk of PTB.^45, 46^ An Irish questionnaire-based COVID-19 study among 71 pregnant women^47^ revealed an increased worry for elderly relatives and children rather than for their pregnancy. Attitudes to the COVID-19 lockdown is being systematically studied as part of the HOPE project^48^ in Denmark, and the feeling of being efficacious, i.e., being able to act on information given, seems to be a very significant predictor of protective behaviour^49^ in western democracies. Surprisingly, the lockdown-associated suspension of daily duties have been shown to be of value for people with vulnerable psyches.^50^ Thus, in Denmark the reduced workplace and social stress may outweigh the negative effects of worries associated with the pandemic. While the end of the lockdown period was associated with improved psychological well-being,^51^ the practicalities of easing lockdown restrictions and returning to work, returning children to school, etc. could increase stress and anxiety. This is particularly true among women with depression (prevalence of 14.2 % among Danish women) or anxiety (prevalence of 6.8 % among Danish women),^52^ both of which have been associated with PTB.^53^ We cannot exclude that the psychosocial and behavioural changes during lockdown, i.e., more people working from home, could have affected the risk of xPTB in specific groups of pregnant women.

The observational nature of this study precludes causal inference, but the sizeable effect on the xPTB rate indicates that it will be worthwhile to identify the elements of the lockdown that have conferred this unintended protective effect regarding xPTB. One might speculate that it will be possible to identify a specific psychosocial phenotype in mid-pregnancy that may benefit from specialised care. However, the complex causality of PTB^30^ suggests that defining controllable protective factors will require a cross-disciplinary effort.

The use of national registers allowed us to examine all Danish pregnancies, thus avoiding selection bias and allowing us to provide a complete assessment of birth and mortality rates.

## Conclusion

The period of COVID-19 restrictions was characterised by a reduction in the extremely preterm live birth rate, in the strict lockdown period, and a reduction in the stillbirth rate throughout the extended period of restrictions. Furthermore, there was a tendency for pregnancies to run to term during this period.

## Data Availability

Data were obtained from Danish Registers and access is possible through contact with Sundhedsdatastyrelsen ("Danish Health Data Authority").

## Acknowledgements

This research was conducted using data obtained from the Danish National registers. The Danish Health Data Authority are acknowledged for their support with preparation and linkage of the data.

## Notes

### Competing Interest Statement

Dr Breindahl has a patent (NeoHelp) with royalties paid. Dr Breindahl has nothing else to disclose. All other authors reported to have nothing to disclose.

### Author Declarations

The study was conducted according to Danish legislation and guidelines for register research and was approved by the Data protection Agency officer at Statens Serum Institut (No:20/04753)

