## Supplemental material for "Preterm birth, stillbirth, and early neonatal mortality during the Danish COVID-19 lockdown"

***Supplementary Material***

***Index:***

*eAppendix	p 1*

Supplement Figure legends	p 2

eFigure 1. Denmark	p 4

eFigure 2. Denmark	p 5

eFigure 3. Netherlands	p 6

eFigure 4. Netherlands	p 7

eFigure 5. USA	p 8

eFigure 6. USA	p 9

eFigure 7. Sweden	p 10

eFigure 8. Sweden	p 11

***eAppendix***

**The timeline and stringency of Danish COVID-19 policies**

The Danish Health Authority first officially oriented the Danish public about the outbreak in Wuhan, China on Jan 8, 2020 (Figure 1).^1^ What followed was a period of close communication with the Danish public (via press releases and conferences) as well as frequent publications of guidelines and recommendations.^1^ On February 27, with the spread of COVID-19 in Asia and Italy throughout February and the diagnosis of the first COVID-19 case in Denmark, the Danish Government issued guidelines on containment, isolation, and surveillance, two days later, COVID-19 was added to the list of contagious diseases covered by the national Epidemics laws (Figure 1).^1^ Revised guidelines, issued Mar 10, expanded the strategy from one of containment to one of control, preventing spread to vulnerable groups and ensuring the treatment capacity in hospitals (Figure 1).^2^

Following a dramatic rise in the number of COVID-19 positive cases (eFigure 1A), and simultaneous WHO declaration of COVID-19 being a pandemic, the Danish government on March 11 announced an extensive nationwide lockdown (Figure 1) involving school closures, work from home regulations, reductions in public transport use, restricted access to hospitals and nursing homes, and limitations on assemblies (eFigure 1B).^1^ The first stage of easing the lockdown was initiated on April 15, we define this period as the “strict lockdown” (March 12, - April 14, 2020). To date, COVID-19 restrictions have continued with varying stringency.

**The Effect of COVID-19 restrictions on mobility**

The strict lockdown period is notably the period with the most stringent control and containment policies (eFigure 1B)^3^. These policies were remarkably effective, as demonstrated by an increase in time spent in places of residence alongside considerable (~50 %) reductions in workplace attendance, visits to retail spaces, and visits to transit stations compared to a baseline value (the median value from the 5-week period Jan 3, - Feb 6, 2020) (eFigures 1C & 2).^4^ The mobility analyses also show that easing of the strict lockdown, on Apr 14, resulted in a steady return to workplaces over the months April, May and June (eFigure 1C), as well as an increase in visits to retail shops and recreational facilities (eFigure 2).

**Supplement Figure legends**

**eFigure 1**. The period of Danish COVID-19 restrictions (time frame plotted: Feb 15, to Sep 30, 2020) under investigation is defined by the graphical representations of **A:** the 7-day rolling mean of cases and deaths, **B:** the stringency index (black line): which is a measure of the strictness of ‘lockdown’ policies with regard to restricting people’s behaviour. The stacked coloured columns represent measures of the containment and control policies implemented across the time frame. Data source for panel A and B: <https://raw.githubusercontent.com/OxCGRT/covid-policy-tracker/master/data/OxCGRT_latest.csv>.^3^ **C:** The daily average mobility of residents of Denmark within workplaces (blue) and places of residence (red) are represented for the time frame. The mobility changes are normalized with respect to a baseline assessment (the median value from the 5-week period January 3, - February 6, 2020) The open circles represent the daily average change from baseline and a very clear shift is noted over the weekends where, thanks to essential workers (nurses, food workers, grocery stores employees, etc.), a much smaller reduction in workplace attendance is noticeable. Data source: Google Mobility Reports.^4^ The lockdown period is demarcated between the vertical dashed-dotted lines.

**eFigure 2**. The daily average mobility of Danish residents. Change in total visitors to retail shops and recreational spaces; groceries and pharmacies (upper panel); parks; transit stations (middle panel) and workplaces (lower panel); as well as the duration of time spent in places of residence (lower panel) are plotted in relation to a baseline value (the median value from the 5-week period January 3, - February 6, 2020). Data source: Google Mobility Reports.^4^ Shaded regions represent the seven-day rolling range.

**eFigure 3** The period of COVID-19 restrictions in the Netherlands (time frame plotted: Feb 15, to Sep 30, 2020) under investigation is defined by the graphical representations of **A:** the 7-day rolling mean of cases and deaths, **B:** the stringency index (black line): which is a measure of the strictness of ‘lockdown’ policies with regard to restricting people’s behaviour. The stacked coloured columns represent measures of the containment and control policies implemented across the time frame. Data source for panel A and B: <https://raw.githubusercontent.com/OxCGRT/covid-policy-tracker/master/data/OxCGRT_latest.csv>.^3^ **C:** The daily average mobility of residents of the Capital Region of Denmark within workplaces (blue) and places of residence (red) are represented for the time frame. The mobility changes are normalized with respect to a baseline assessment (the median value from the 5-week period January 3, - February 6, 2020) The open circles represent the daily average change from baseline and a very clear shift is noted over the weekends where, thanks to essential workers (nurses, food workers, grocery stores employees, etc.), a much smaller reduction in workplace attendance is noticeable. Data source: Google Mobility Reports.^4^ The lockdown period is demarcated between the vertical dashed-dotted lines.

**eFigure 4**. The daily average mobility of residents in the Netherlands. Change in total visitors to retail shops and recreational spaces; groceries and pharmacies (upper panel); parks; transit stations (middle panel) and workplaces (lower panel); as well as the duration of time spent in places of residence are plotted in relation to a baseline value (the median value from the 5-week period January 3, - February 6, 2020). Data source: Google Mobility Reports.^4^ Shaded regions represent the seven-day rolling range.

**eFigure 5** The period of COVID-19 restrictions in the US (time frame plotted: Feb 15, to Sep 30, 2020) under investigation is defined by the graphical representations of **A:** the 7-day rolling mean of cases and deaths, **B:** the stringency index (black line): which is a measure of the strictness of ‘lockdown’ policies with regard to restricting people’s behaviour. The stacked coloured columns represent measures of the containment and control policies implemented across the time frame. The grey shaded areas represent the range of start and end dates of lockdowns across the states. Data source for panel A and B: <https://raw.githubusercontent.com/OxCGRT/covid-policy-tracker/master/data/OxCGRT_latest.csv>.^3^ **C:** The daily average mobility of residents of the Capital Region of Denmark within workplaces (blue) and places of residence (red) are represented for the time frame. The mobility changes are normalized with respect to a baseline assessment (the median value from the 5-week period January 3, - February 6, 2020) The open circles represent the daily average change from baseline and a very clear shift is noted over the weekends where, thanks to essential workers (nurses, food workers, grocery stores employees, etc.), a much smaller reduction in workplace attendance is noticeable. Data source: Google Mobility Reports.^4^ The first state-wide lockdown initiation date and last state-wide lockdown easing date are indicated as vertical dashed-dotted lines.

**eFigure 6**. The daily average mobility of residents in the US. Change in total visitors to retail shops and recreational spaces; groceries and pharmacies (upper panel); parks; transit stations (middle panel) and workplaces (lower panel); as well as the duration of time spent in places of residence are plotted in relation to a baseline value (the median value from the 5-week period January 3, - February 6, 2020). Data source: Google Mobility Reports.^4^ Shaded regions represent the seven-day rolling range.

**eFigure 7.** The period of Swedish COVID-19 restrictions (time frame plotted: Feb 15, to Sep 30, 2020) under investigation is defined by the graphical representations of **A:** the 7-day rolling mean of cases and deaths, **B:** the stringency index (black line): which is a measure of the strictness of ‘lockdown’ policies with regard to restricting people’s behaviour. The stacked coloured columns represent measures of the containment and control policies implemented across the time frame. Data source for panel A and B: <https://raw.githubusercontent.com/OxCGRT/covid-policy-tracker/master/data/OxCGRT_latest.csv>.^3^ **C:** The daily average mobility of residents of Sweden within workplaces (blue) and places of residence (red) are represented for the time frame. The mobility changes are normalized with respect to a baseline assessment (the median value from the 5-week period January 3, - February 6, 2020) The open circles represent the daily average change from baseline and a very clear shift is noted over the weekends where, thanks to essential workers (nurses, food workers, grocery stores employees, etc.), a much smaller reduction in workplace attendance is noticeable. Data source: Google Mobility Reports.^4^ The lockdown period is demarcated between the vertical dashed-dotted lines.

**eFigure 8**. The daily average mobility of Swedish residents. Change in total visitors to retail shops and recreational spaces; groceries and pharmacies (upper panel); parks; transit stations (middle panel) and workplaces (lower panel); as well as the duration of time spent in places of residence are plotted in relation to a baseline value (the median value from the 5-week period January 3, - February 6, 2020). Data source: Google Mobility Reports.^4^ Shaded regions represent the seven-day rolling range.

**eFigure 1. Denmark**

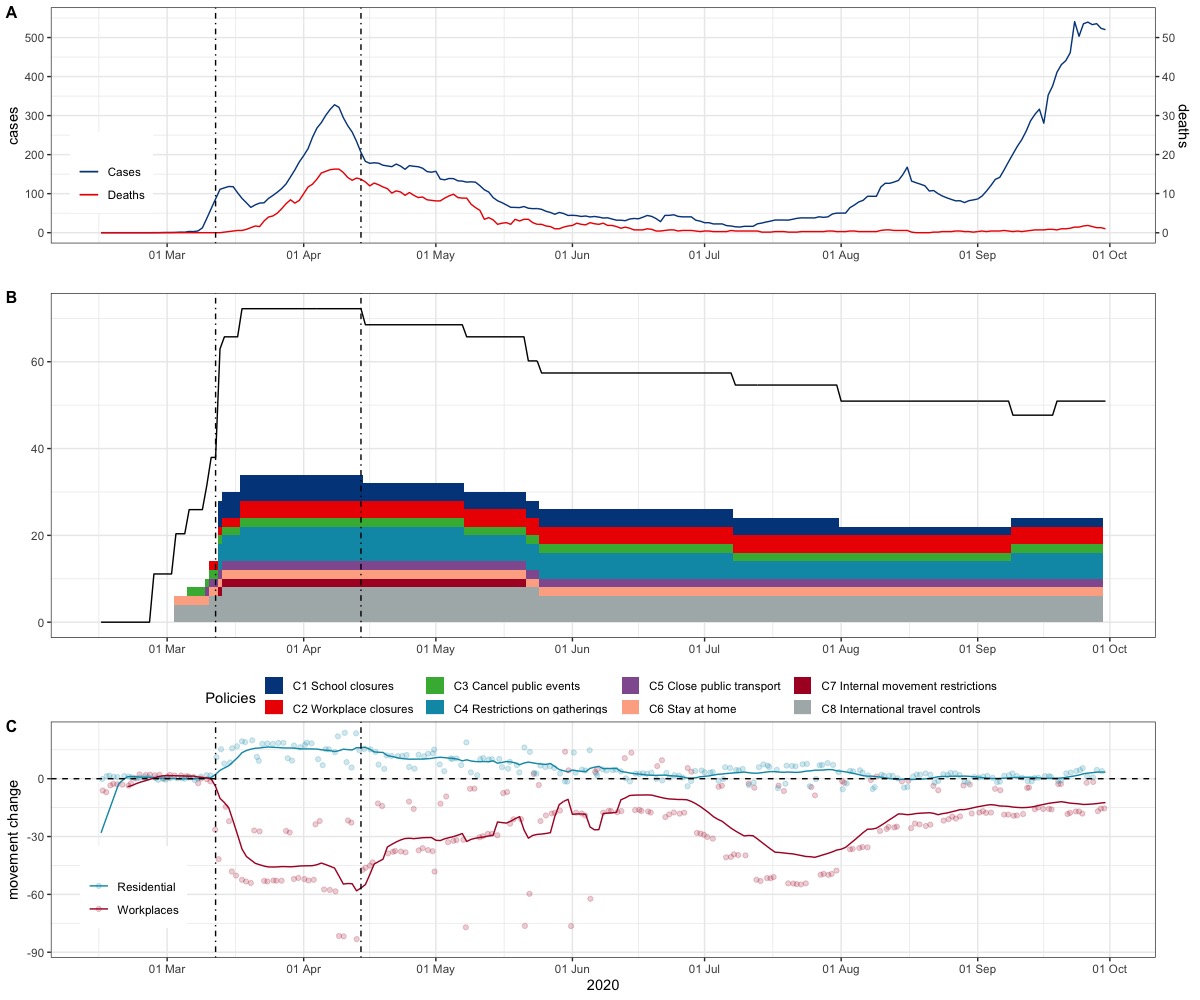

**eFigure 2. Denmark**

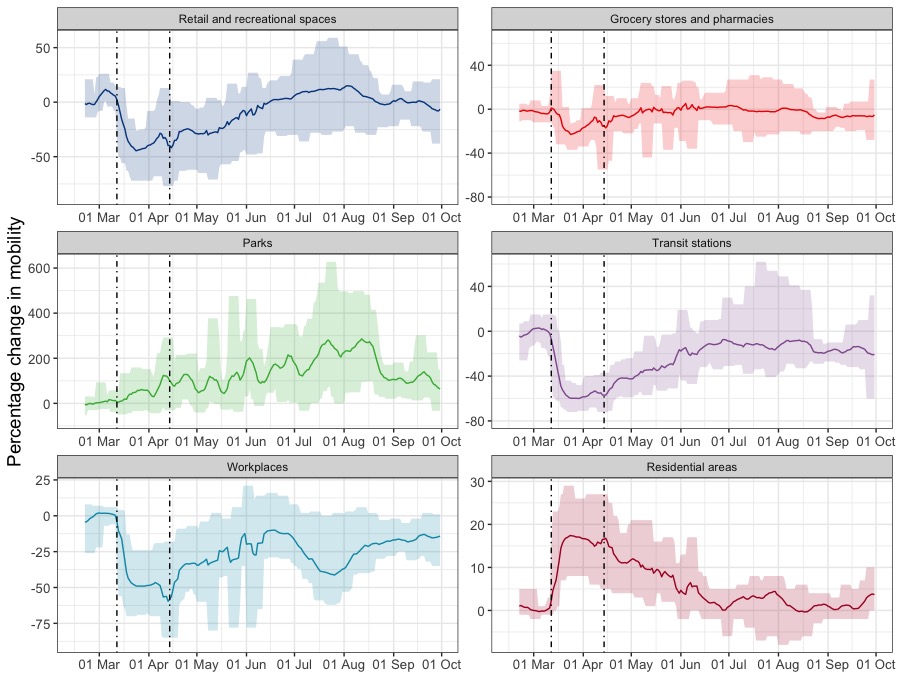

**eFigure 3. Netherlands**

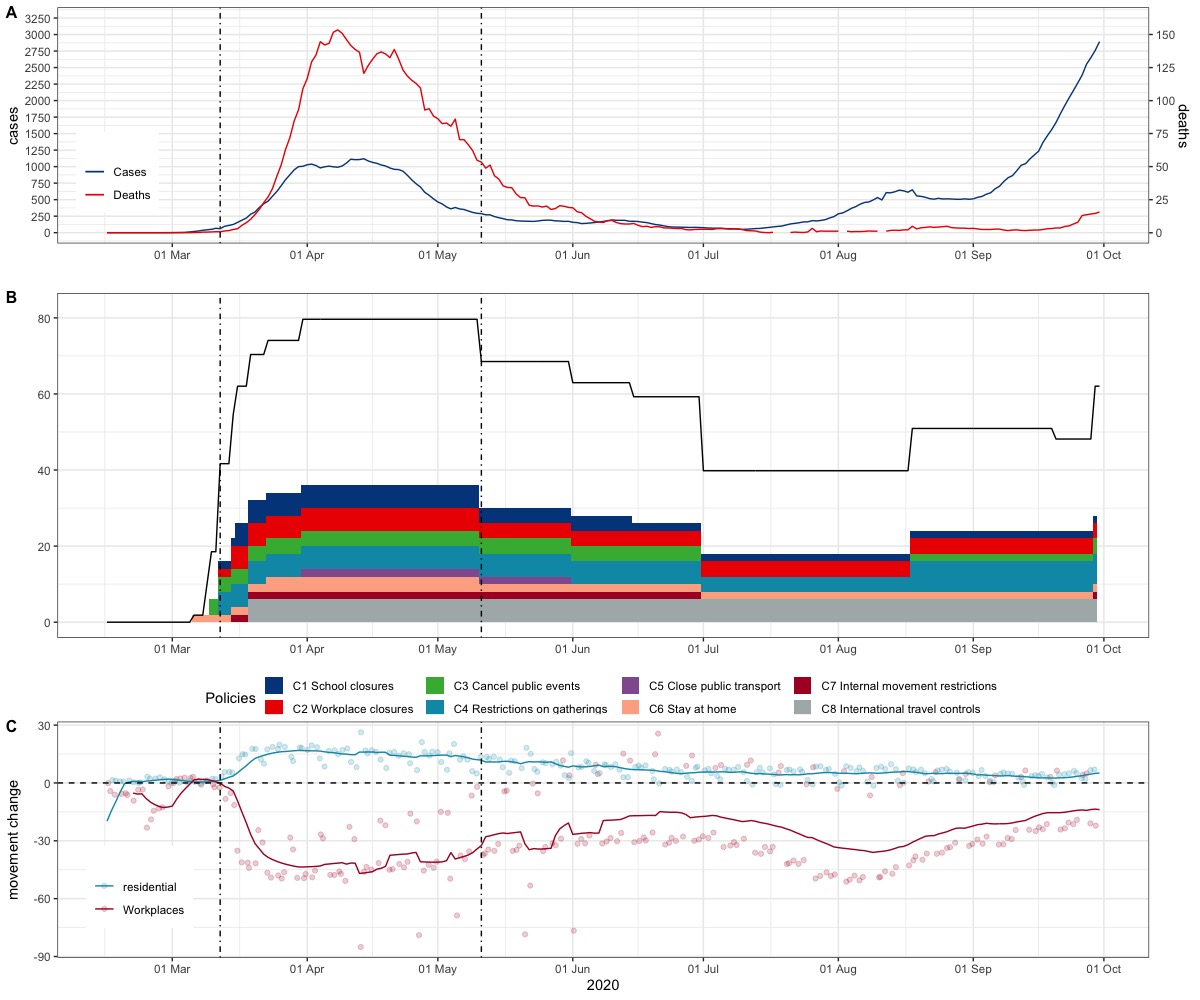

**eFigure 4. Netherlands**

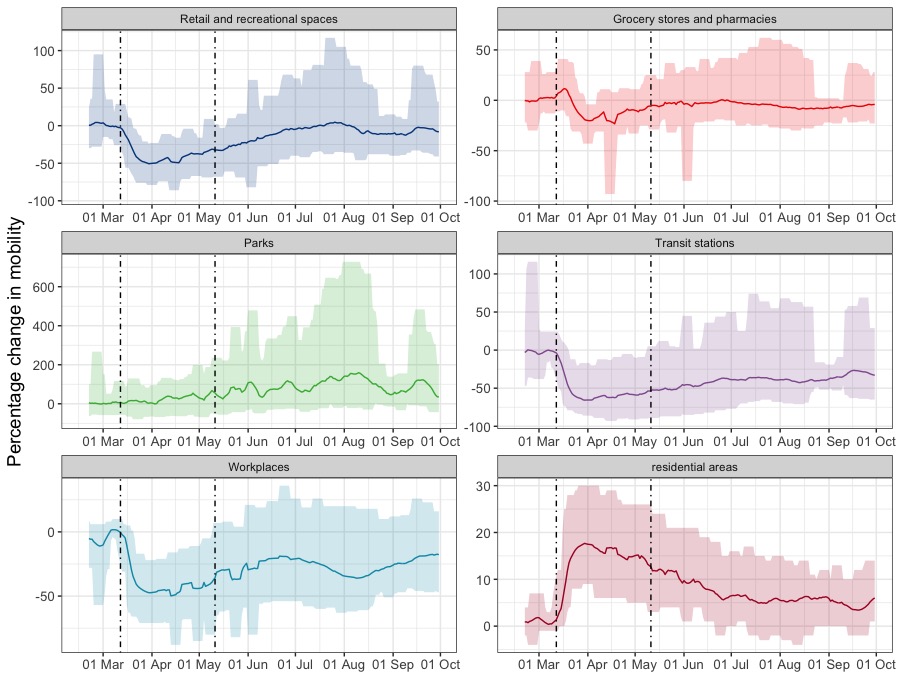

**eFigure 5. USA**

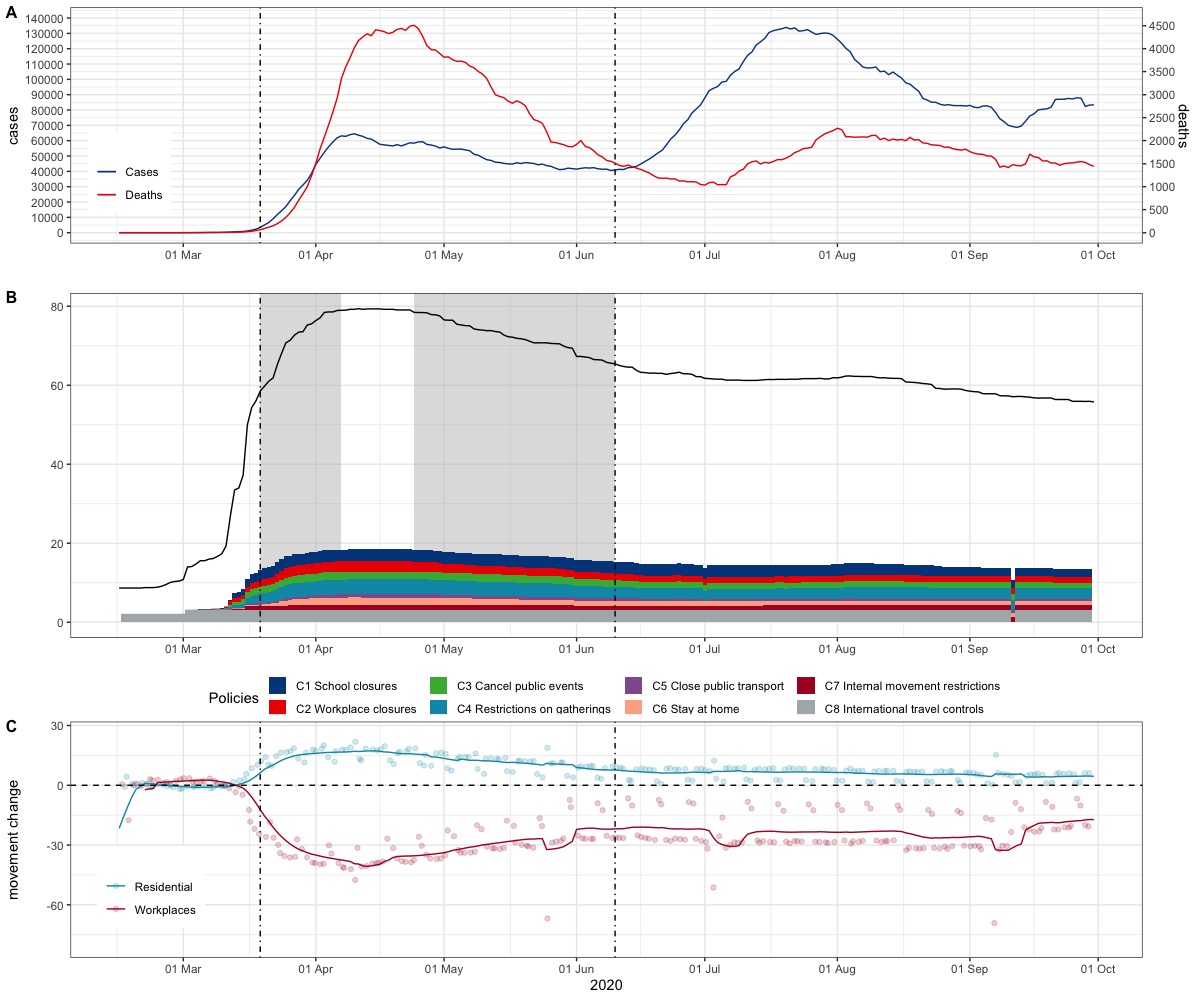

**eFigure 6. USA**

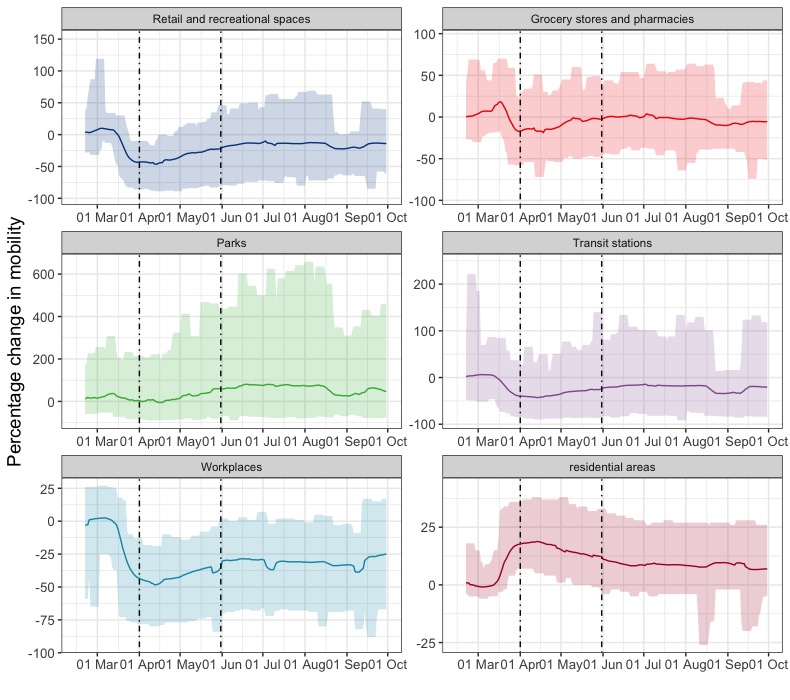

**eFigure 7. Sweden**

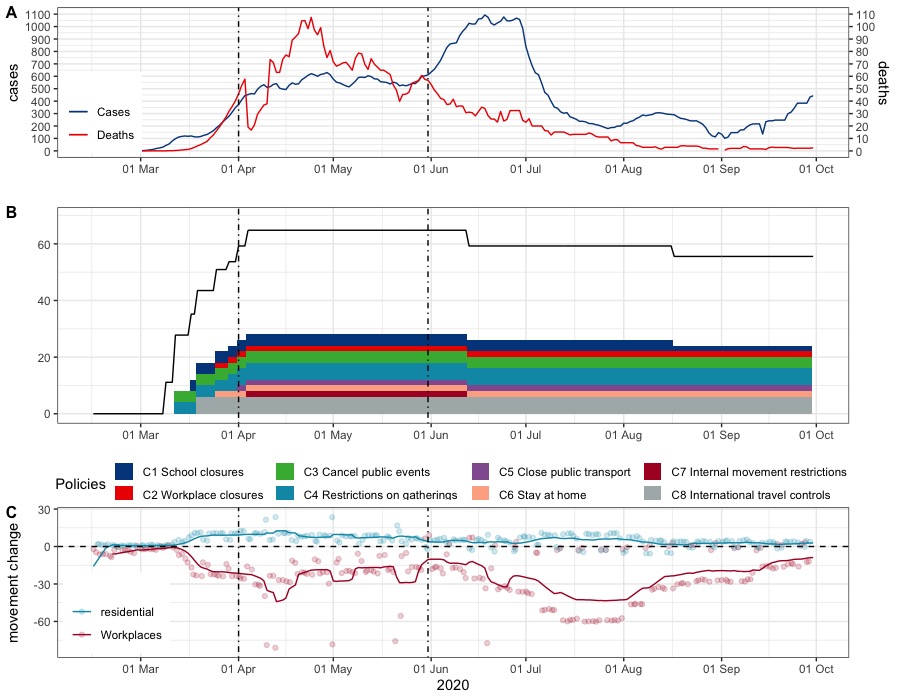

**eFigure 8. Sweden**

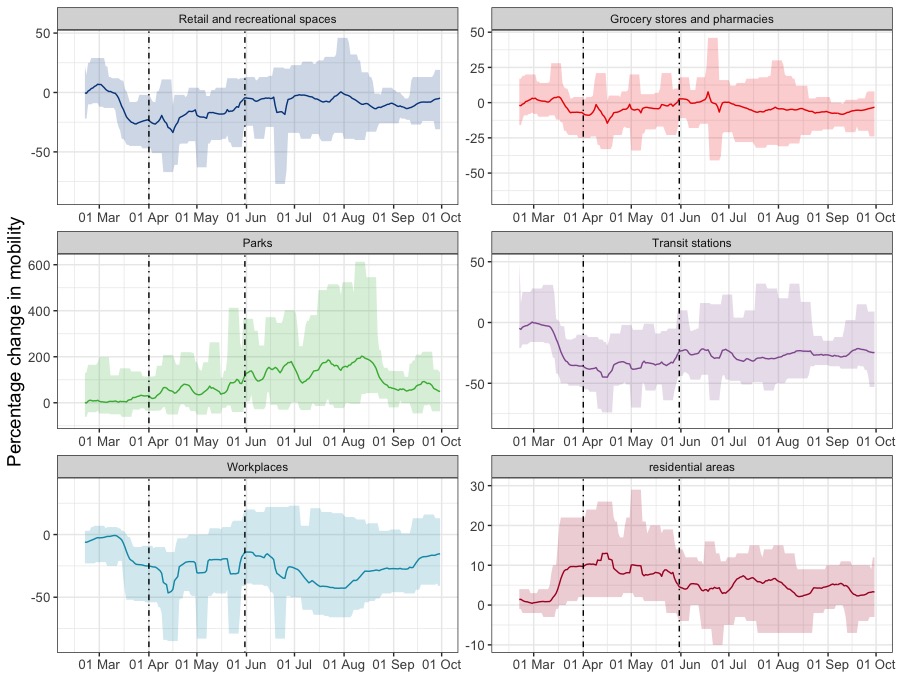
